# Categories of intimate partner violence and abuse (IPVA) among young women and men: Latent Class Analysis of psychological, physical, and sexual victimisation and perpetration in a UK birth cohort

**DOI:** 10.1101/2021.08.20.21262361

**Authors:** Annie Herbert, Abigail Fraser, Laura D. Howe, Eszter Szilassy, Maria Barnes, Gene Feder, Christine Barter, Jon Heron

## Abstract

**Background:** In the UK, around one-third of young people are exposed to IPVA by 21 years old. However, types of IPVA victimisation in this population (psychological, physical, sexual), and their relationship with impact and perpetration are poorly understood.

**Methods:** Participants in a UK birth cohort reported IPVA victimisation and perpetration by age 21. We carried out a latent class analysis, where we categorised IPVA by types/frequency of victimisation, and then assigned individuals to their most probable class. Within these classes, we then estimated risks of reported: 1) types of negative impacts (sad, upset/unhappy, anxious, depressed, affected work/studies, angry/annoyed, drank/took drugs more); 2) types/frequency of perpetration.

**Results:** Among 2,130 women and 1,149 men, 32% and 24% reported IPVA victimisation (of which 89% and 73% reported negative impact); 21% and 16% perpetration. Victimisation responses were well represented by five classes, including three apparent in both sexes: *No-low victimisation* (characterised by low probabilities of all types of victimisation; average probabilities of women and men belonging to this class were 82% and 70%); *Mainly psychological* (15% and 12%); *Psychological & physical victimisation* (4% and 7%), and two classes that were specific to women: *Psychological & sexual* (7%); *Multi-victimisation* (frequent victimisation for all three types; 4%). In women, all types of negative impact were most common in the *Psychological & sexual* and *Multi-victimisation* classes; for men, the *Psychological & physical* class. In women, all types of perpetration were most common for the *Mainly psychological, Psychological & physical*, and *Multi-victimisation* classes; in men, the *Mainly psychological* and *Psychological & physical* classes.

**Discussion:** In this study of young people, we found categories of co-occurrence of types and frequency of IPVA victimisation associated with differential risks of negative impact and perpetrating IPVA. This is consistent with emerging evidence of IPVA differentiation and its variable impact in other populations.

## Background

Among young people in the UK, it is estimated that one-third to three-quarters are exposed to Interpersonal Violence and Abuse (IPVA) victimisation by 21 years old, and one fifth perpetrate IPVA (Barter, 2009b; Herbert et al., 2020; Young et al., 2019; Young et al., 2018). Evidence mainly from north America suggests poor mental and physical outcomes in young people who experience IPVA, and so effective interventions for its prevention and related outcomes are needed (Barter & Stanley, 2016; J. Campbell et al., 2002).

Yet patterns of IPVA victimisation in the UK context, and their relationship to perpetration are poorly understood. Researchers have measured the prevalence of IPVA either as a binary concept, or of different types such as psychological or physical IPVA alone (Barter, 2009b; Herbert et al., 2020; Young et al., 2019; Young et al., 2018), but we do not yet know the extent to which different types of IPVA co-occur for the same individual. Understanding co-occurrence and the potential differential impact of different combinations of abuse is crucial for developing interventions to support people experiencing IPVA and for its prevention. It has been argued that severity, frequency, and impact of IPVA are important aspects of IPVA to consider, particularly in exploring IPVA by sex (Bacchus, Buller, Ferrari, Brzank, & Feder, 2018; Hegarty et al., 2012; Hester, Donovan, & Fahmy, 2010; Walby & Allen, 2004; Walby & Towers, 2018). For example, prevalence of psychological IPVA may not be ‘gendered’, but the potential health damage of continuous coercive control is (Finkelhor, Ormrod, & Turner, 2007; Herbert et al., 2020; Walby & Towers, 2018). A more differentiated categorisation of IPVA victimisation and perpetration, considering types, severity, frequency, and impact, is needed to evaluate its impact on individuals and families. Finally, prevalence studies in young people have either addressed victimisation alone, or presented estimates for victimisation and perpetration separately (Barter, 2009b; Herbert et al., 2020; Young et al., 2019; Young et al., 2018), but it is important to understand to what extent IPVA is inflicted by both partners within a romantic relationship (Herbert et al., 2020; Miller et al., 2013; O’Leary & Slep, 2012).

Against this background, we investigated patterns of IPVA victimisation experiences among young people aged up to 21 in a large UK population-based birth cohort according to types of abuse (psychological, physical, sexual), severity (e.g., coercive vs. forced sexual), and frequency. We then explored relationships between these patterns and different types of self-reported negative impact and IPVA perpetration. We aimed to provide a better understanding of patterns of different types, frequency, and impacts of IPVA among young men and women in the UK.

## Methods

### Participants and data collection

We analysed data on 3,279 young people who were part of the ALSPAC (Avon Longitudinal Study of Parents and Children, formerly ‘Children of the 90s’) birth cohort study, and who had answered questions relating to IPVA (delivered in online and paper form) at 21 years old. Demographic, behavioural, and health characteristics of this sample have been described previously (Herbert et al., 2020; A. Yakubovich, Heron, Feder, Fraser, & Humphreys, 2019). Briefly, two-thirds were female, with a median age of 21 years (interquartile range: 21 to 22), the large majority were white, self-defined as heterosexual, with 92% indicating that they had been in a relationship by age 21 (Herbert et al., 2020).

ALSPAC recruited pregnant women resident in Avon, UK, with expected dates of delivery 1st April 1991 to 31st December 1992, as well as their partners and offspring (our analyses focus on the offspring at age 21). The initial number of pregnancies enrolled was 14,541, resulting in 13,988 children who were alive at 1 year of age. When the oldest children were approximately 7 years of age, an attempt was made to bolster the initial sample with eligible cases who had failed to join the study originally – resulting in 15,454 total enrolled pregnancies, of which 14,901 foetuses were alive at 1 year of age. More information on ALSPAC data is available in published cohort profiles (Boyd et al., 2013; Fraser et al., 2013; Northstone et al., 2019), and the study website, which contains details of all the data available through a fully searchable data dictionary and variable search tool (http://www.bristol.ac.uk/alspac/researchers/our-data/).(“Avon Longitudinal Study of Parents and Children. Explore data and samples.,”)

Our analysis uses data from the age 21 questionnaire, which was distributed online in mid-December 2013, followed by a series of up to four reminders at three-week intervals, some of these reminders containing a paper version of the same questionnaire. 9,353 participants were sent a questionnaire (online and paper options), to which 37% (n=3,459) responded. The current study’s cohort was the 3,279 who answered questions within the IPVA section (minus one participant where sex was missing).

The IPVA section of the questionnaire at age 21 was developed by IPVA researchers, based on previous UK and European questionnaires and the PROVIDE questionnaire (Barter, 2009b; Hester et al., 2015), and is described in full in a paper validating its psychometric properties (A. Yakubovich et al., 2019). The questions on IPVA were approved by the ALSPAC Ethics and Law Committee (ref: E201210).

Study data were collected and managed using REDCap electronic data capture tools hosted at University of Bristol (Harris et al., 2019). Ethical approval for the study was obtained from the ALSPAC Ethics and Law Committee and the Local Research Ethics Committees. Full details of the ALSPAC consent procedures are available on the <u>study website</u> (http://www.bristol.ac.uk/alspac/researchers/research-ethics/).

### IPVA victimisation measures

Within the IPVA section of the questionnaire, participants were asked “How often altogether have any of your partners ever done any of the following to you and how old were you?”, with “by ‘partner’, we mean anyone you have ever been out with or had a relationship with, long-term or short-term (including ‘one-night stands’)”. The eight different examples of victimisation that followed are presented in **Table 1**, along with short-hand labels that we employ in the current study.

**Table 1.** Intimate Partner Violence and Abuse (IPVA) question wording, and labelling employed in the current study

|  |  |
| --- | --- |
| Victimisation |  |
| Question wording | Type label in current study |
| “Told you who you could see and where you could go and/or regularly | Coercive psychological |
| “Made fun of you, called your hurtful names, shouted at you?” | Explicit psychological |
| “Used physical force such as pushing, slapping, hitting or holding you | Physical 1 |
| “Used more severe physical force such as punching, strangling, beating you | Physical 2 |
| “Pressured you into kissing/touching/e) something else?” | Coercive sexual 1 |
| “Physically forced you into kissing/touching/something | Coercive sexual 2 |
| “Pressured you into having sexual intercourse?” | Forced sexual 1 |
| “Physically forced you into having sexual intercourse?” | Forced sexual 2 |
| Perpetration |  |
| Question wording | Type label in current study |
| “Told them who they could see and where they could go and/or regularly | Coercive psychological |
| “Made fun of them, called them hurtful names, shouted at them?” | Explicit psychological |
| “Hit, slapped, kicked or otherwise physically hurt them?” | Physical |
| “Pressured or forced them into kissing, touching, sexual intercourse or any other sexual activity when they did not want to?” | Sexual |

We employed the labels *Physical 1* and *Physical 2, Coercive sexual 1* and *Coercive sexual 2*, and *Forced sexual 1* and *Forced sexual 2*, where we considered them to represent different severities (2 being higher than 1) of physical, coercive sexual, and forced sexual IPVA, respectively. We did not consider *Coercive psychological* or *Forced psychological* to have a particular order in terms of severity between each other. We made no assumptions about ordering of severity when modelling the above eight variables in Latent Class Analysis (described later under ‘Statistical Analysis’), the purpose of the labelling is to aid with interpretation, only.

The options for frequency of the above eight events were: ‘never’, ‘once’, ‘a few times’, ‘often’. We treated these variables as ordinal categorical responses, where ‘a few times’ and ‘often’ were combined into one category (given small numbers of endorsements to ‘often’). For men, numbers of endorsements to the two questions relating to *Coercive sexual 2* and *Forced sexual 2* (i.e., relating to intercourse) were considered too small for these data to be included in analyses (19 men reported *Coercive Sexual 2* either ‘a few times’ or ‘often’, 6 for Forced Sexual 2). Therefore, although all eight victimisation questions were included in analyses for women, these two sexual victimisation questions were not considered any further in analyses for men (leaving the six remaining victimisation questions).

### Impact of IPVA victimisation

Following the eight victimisation questions, participants were asked: “How did you feel after they did these things to you?” with the following examples of negative impact given: ‘upset/unhappy’; ‘affected my work/studies’; ‘made me feel sad’; ‘anxious’; ‘made me drink more alcohol/take more drugs’; ‘angry/annoyed’; ‘depressed’. Each of these impact responses were binary yes/no variables.

### IPVA perpetration measures

Participants were then asked: “How often altogether have you done any of the following to any of your partners, and how old were you?”. The four different examples of perpetration (similarly worded to the examples of victimisation, but due to questionnaire space, slightly more condensed) are presented in **Table 1**.

### Statistical analyses

We carried out all analyses separately for women and men. A large part of the literature has focussed solely on violence against women and so sex-stratified analyses would allow comparison with existing literature (J. C. Campbell, 2002; Ellsberg, Jansen, Heise, Watts, & Garcia-Moreno, 2008; Garcia-Moreno et al., 2013; Vezina & Hebert, 2007; A. R. Yakubovich et al., 2018); also small numbers led us to only be able to analyse six of the eight victimisation questions for men.

We carried out a bias-adjusted three-step latent class analysis (LCA) (Heron, Croudace, Barker, & Tilling, 2015; Vermunt, 2010). First, we carried out an LCA where the eight victimisation questions (six for men; as ordinal variables ‘Never’, ‘Once’, ‘A few times+’) were considered. An LCA involves fitting the data to a pre-assigned number of classes using a multivariate mixture model. Within each class there is an estimated probability per response variable (e.g., if the variables included were binary: the first class may represent a high probability of explicit psychological victimisation, low probability of physical, etc., with the second class being characterised by a different set of probabilities for each of the victimisation measures). Each individual in the analysis then has a posterior probability of belonging to each class, based on their observed responses. Then, we decided on the final class solution based on different indicators of goodness-of-fit, face validity, a possible sex-invariant solution, and utility (classes that would not be too small). We checked the entropy of the class solution to determine how reliably individuals could be assigned to the different classes in the solution. Further details of this process are provided in **Supplementary Box S1**.

In the second step, we assigned each of the study participants to one of the derived classes based on their modal posterior probability of membership.

Finally, in the third step, to determine whether impact and perpetration varied between different classes, we fitted a separate logistic regression model for different types of negative impact, and different types of perpetration as the dependent variable (outcome), respectively (all binary outcomes; in the case of perpetration outcomes, at least ‘Once’ vs. ‘Never’). Class assignment was included as an independent variable (exposure), with a correction (bias-adjustment) for potential misclassification error (Heron et al., 2015; Vermunt, 2010).

The study-specific three-step process is described in more detail in **Supplementary Box S1**. We prepared all data in Stata version 15.1, ran LCAs in Mplus version 8.4, and derived proportions of impact and perpetration types and created plots using R version 3.5.1. The Mplus and R scripts used for analyses are available at: https://github.com/pachucasunrise/IPVA_categories.

## Results

Of 2,130 women and 1,149 men who responded to questionnaires at age 21, 880 (41%) and 330 (29%) reported IPVA victimisation, respectively. The most common victimisation type was psychological (35% and 26%), followed by physical (18% and 10%), then sexual (18% and 5%) (Herbert et al., 2020). Among those reporting IPVA victimisation, 89% of women and 72% of men and reported any negative impact. Of the 2,130 women and 1,149 men, 25% and 20% reported perpetration, respectively, the most common types again being psychological, followed by physical, then sexual.

### IPVA victimisation profiles

**Supplementary Box S1 and Table S1** provide detail on decisions made and model diagnostics between different class solutions. Briefly, indicators were inconsistent in that they pointed to the optimal solution being two or three classes in men, and four, five, or more than six classes in women. Based on choosing the highest number of classes per sex to avoid missing important variation, face validity, and avoiding classes of small numbers, we chose to present the three and five -class solutions for men and women, respectively.

The classes are presented in **Figure 1**. Classes 1-3 in the five-class solution in women were similar in terms of patterns of victimisation types and frequency to Classes 1-3 of the three-class solution in men. However, note that these classes – in terms of exact probabilities of each of the types of victimisation and their frequencies – though similar are not necessarily the same between sexes, as they were derived from separate models.

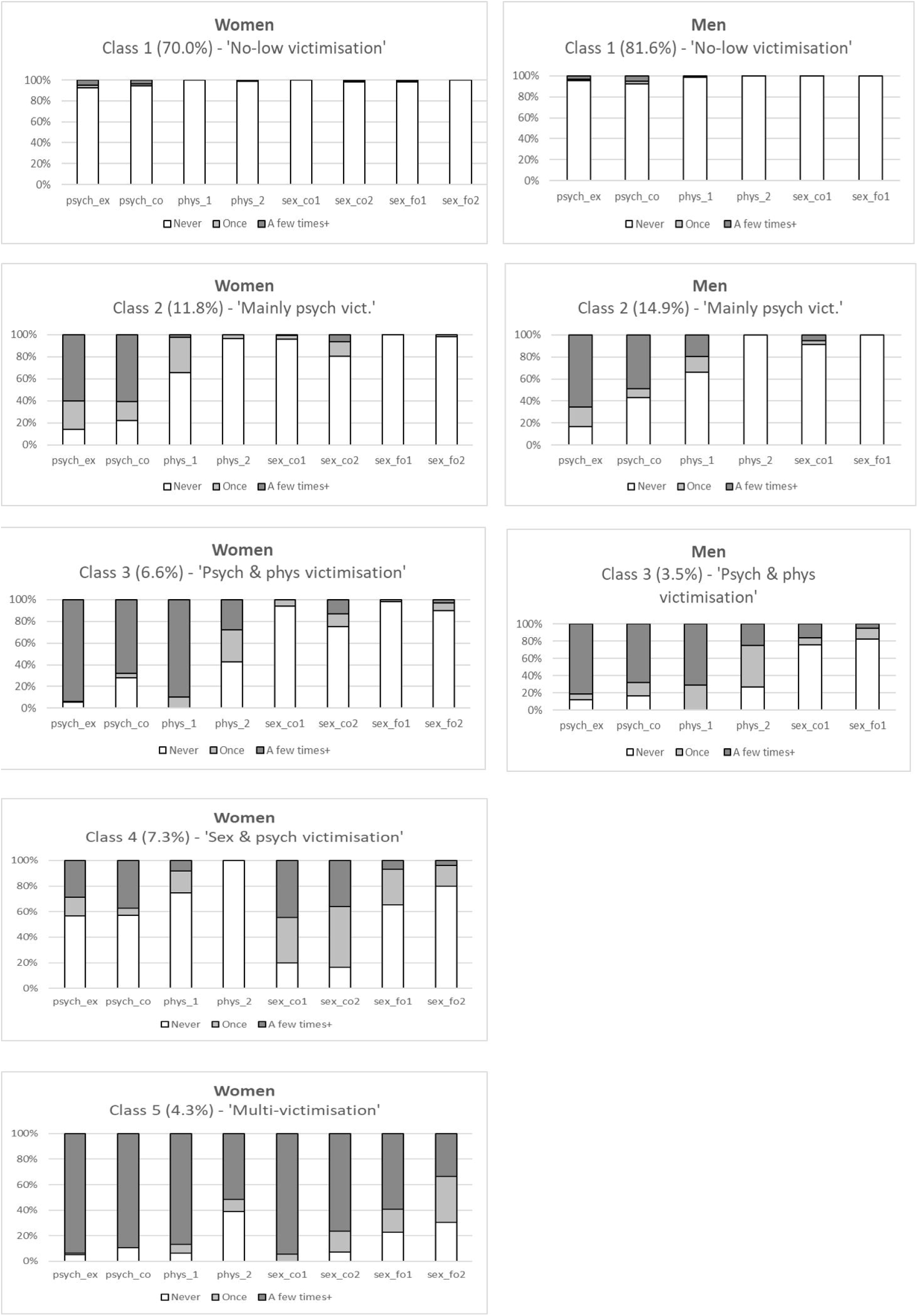

The classes were as follows:

Class 1: *Low-no victimisation* (average posterior probabilities of belonging to this class were high (70% for women and 82% for men). This class was characterised by small chances (0-5%) of coercive or explicit psychological, physical, or sexual victimisation.

Class 2: *Mainly psychological victimisation* (average probabilities of belonging to this class, women: 12%, men: 15%). This class was characterised by frequent psychological victimisation. Within this class there was an over 50% chance of frequent (i.e. at least ‘a few times’) explicit or coercive psychological victimisation. At least 30% chance of experiencing physical victimisation of the type ‘Physical 1’ (i.e. ‘such as pushing, slapping, hitting or holding you down’).

Class 3: *Psychological & physical victimisation* (women: 7%, men: 4%). Characterised by frequent psychological and physical victimisation. Within this class there was an over 60% chance of frequent (i.e., at least ‘a few times’) explicit or coercive psychological victimisation and of frequent physical victimisation of the type *Physical 1* (i.e., ‘such as pushing, slapping, hitting or holding you down’). Over 50% chance of reporting at least ‘Once’ for *Physical 2* (i.e., ‘such as punching, strangling, beating you up, hitting you with an object’), which was more likely to occur ‘Once’ compared to ‘A few times+’.

Classes 4 and 5 related to women only:

Class 4: *Psychological & sexual victimisation* (7%). Characterised by coercive sexual victimisation, with a 36-48% chance of reporting frequent (‘A few times+’; depending on whether type 1 or 2), and 35-48% ‘Once’. This class was associated with over 40% chance of reporting either explicit or coercive psychological victimisation, most of this being ‘A few times+’.

Class 5: *Multi-victimisation* (4%). This class was defined by very high chances (>80%) of being explicitly or coercively psychologically victimised, physically victimised of the type *Physical 1* (i.e., ‘such as pushing, slapping, hitting or holding you down’), or coercively sexually victimised. There were also high chances (>50%) of being physically victimised of the type *Physical* 2 (i.e., ‘such as punching, strangling, beating you up, hitting you with an object’) or forcefully sexually victimised. In this class probabilities of being victimised frequently (‘A few times+’) were always much higher than ‘Once’.

Entropy was relatively high: 0.90 for the five-class solution in women and 0.86 for the three-class solution in men.

### IPVA victimisation impact outcomes

After assigning individuals to classes based on their modal posterior probability, the risks of different impact types by the five classes of women and three classes of men were as shown in **Figure 2** (exact values for risks and standard errors are presented in **Supplementary Table S2**). Between the five classes in women, there was a general pattern of being lowest in the *No-low victimisation* class (class 1), increasing in the *Mainly psychological victimisation* class (class 2), then *Psychological & physical victimisation* class (class 3), *Psychological & sexual victimisation* class (class 4), and finally the *Multi-victimisation* class (class 5) (noting the increased surface area covered by each class in **Figure 2**). In men, risks of each type of negative impact were lowest in the *No-low victimisation* class (class 1), increased in the *Mainly psychological victimisation* class (class 2), followed a further increase in the *Psychological & physical victimisation* class (class 3).

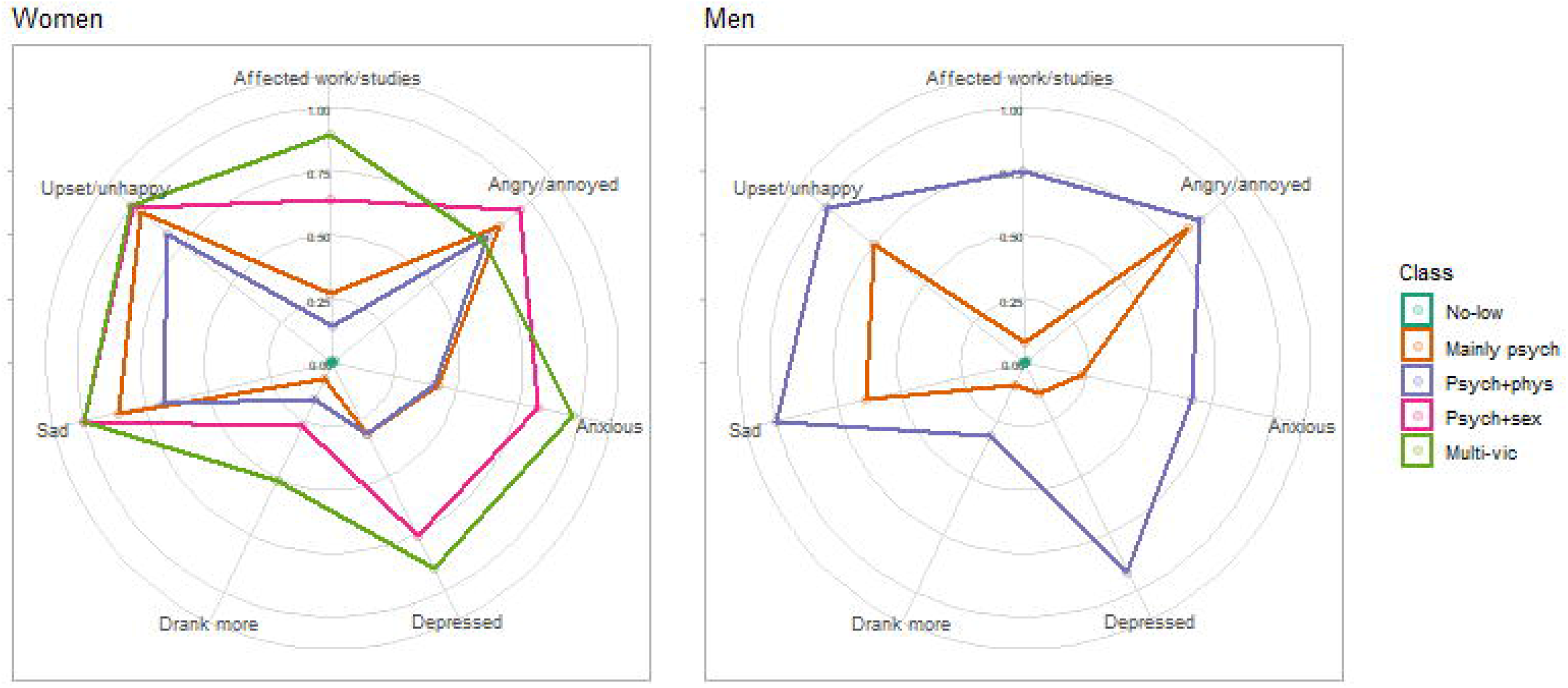

Within each class and regardless of sex, the risks of negative impact tended to be highest for ‘Angry/annoyed’, ‘Upset/unhappy’, and ‘Sad’, and the lowest for ‘Made me drink more alcohol/take more drugs’.

### IPVA perpetration outcomes

The risks of different perpetration types by victimisation classes are shown in **Figure 3** (exact values for risks and standard errors are presented in **Supplementary Table S3**).

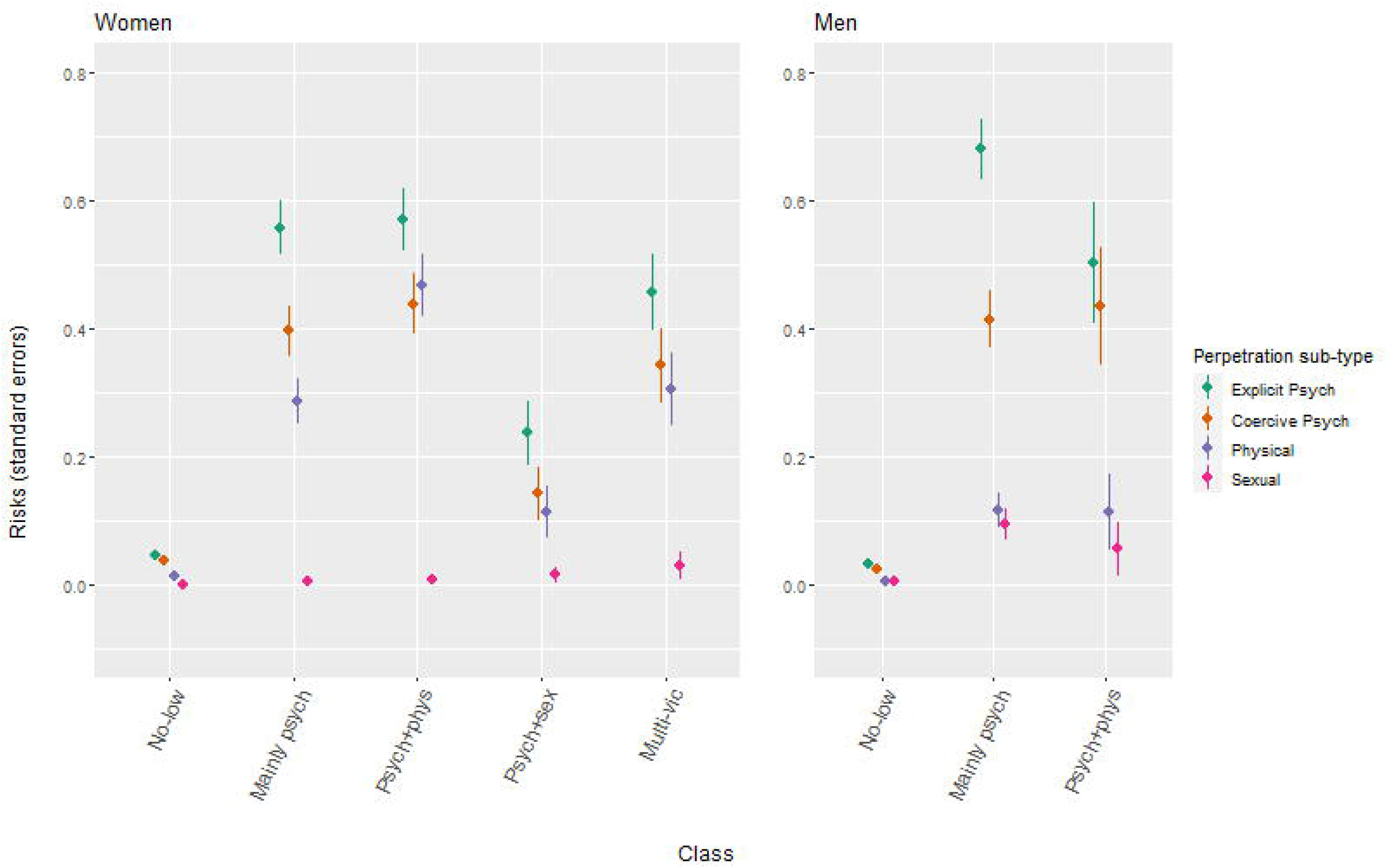

In women, risks of explicit psychological, coercive psychological, and physical perpetration, were generally highest in the *Mainly psychological victimisation* class (class 2), *Psychological & physical victimisation* class (class 3), and *Multi-victimisation* class (class 5), and lowest in the *No-low victimisation* class (class 1). Risks of sexual perpetration were extremely low for all classes in women.

Risks of different perpetration types were higher in the *Mainly psychological victimisation* class (class 2) and *Psychological & physical victimisation* class (class 3) when compared with the *No-low victimisation* class (class 1). There were no notable differences in risks between the *Mainly psychological victimisation* class (class 2) and *Psychological & physical victimisation* class (class 3), except that the risk of explicit psychological perpetration was higher in the *Mainly psychological victimisation* class (class 2).

Within each class and regardless of sex, the risks of different perpetration types were highest for explicit psychological perpetration and the lowest for sexual perpetration.

## Discussion

This is one of few contemporary studies, and the first UK one, to articulate categories of co-occurrence of IPVA victimisation types (psychological, physical, and sexual) and frequency among young people, and to address co-occurrence of victimisation with either negative impact and/or perpetration. We identified five classes of IPVA victimisation types and frequency, two of which only applied to women. There were differential risks of negative impact and of IPVA perpetration types, according to these classes.

There are multiple IPVA studies of young people that have used LCA, but these studies almost exclusively come from north America, whereby the social and educational context greatly differs, as do modes of violence (e.g. the prevalence of violence through gun crime), compared with in the UK (Brooks-Russell, Foshee, & Ennett, 2013; French, Bi, Latimore, Klemp, & Butler, 2014; Goncy, Sullivan, Farrell, Mehari, & Garthe, 2017; Haynie et al., 2013; Hebert, Moreau, Blais, Oussaid, & Lavoie, 2018; Lapierre, Paradis, Todorov, Blais, & Hebert, 2019; Martin-Storey & Fromme, 2016; Mumford, Liu, & Taylor, 2019; Sessarego, Siller, & Edwards, 2019; Swartout, Cook, & White, 2012; Weir & Kaukinen, 2019). In addition, the broad majority of these LCA studies address longitudinal patterns/trajectories of either IPVA victimisation or IPVA perpetration occurrence over time, but not co-occurrence of different victimisation or perpetration types.(Brooks-Russell et al., 2013; Lapierre et al., 2019; Martin-Storey & Fromme, 2016; Miller et al., 2013; Mumford et al., 2019; Swartout et al., 2012; Weir & Kaukinen, 2019)

North American school-based LCA studies addressing co-occurrence of different IPVA types have reported mixed results (Goncy et al., 2017; Haynie et al., 2013; Hebert et al., 2018; Sessarego et al., 2019). First, class characteristics appeared to differ by age. Haynie *et al* and Goncy *et al* studied younger adolescents in middle school (mean ages 13 in each sample) (Goncy et al., 2017; Haynie et al., 2013). Both studies analysed victimisation and perpetration items simultaneously (i.e., within the same LCA, where the model does not distinguish between victimisation items and perpetration items) and did not include any items regarding sexual violence or abuse in their analyses. Though Haynie *et al* reported three classes, and Goncy *et al* five, both reported classes that distinguished *no-low victimisation or perpetration, psychological victimisation or perpetration only*, and *physical victimisation or perpetration only*. Hébert *et al* and Sessarego *et al* studied older adolescents from high-school (mean ages 15 in each sample), and reported three- and four-class solutions, respectively (Hebert et al., 2018; Sessarego et al., 2019). Both reported some form of *no-low victimisation* class (Hébert *et al* reporting separate *no victimisation* and *low victimisation* classes), and a *multi-victimisation* class that was characterised by high probabilities of psychological, physical, and sexual victimisation. Sessarego *et al* also reported a *sexual & psychological victimisation* class.

Of the four studies, Haynie *et al* and Hébert *et al* fitted models separately by sex, and found similar class solutions between the sexes. An exception is that Hébert *et al* found that the form of sexual victimisation in the sexual and psychological victimisation class differed between girls and boys (the former more likely responding attempted/completed rape and the latter responding unwanted sexual contacts) (Haynie et al., 2013; Hebert et al., 2018).

Three studies explored other outcomes in their class solutions. Haynie *et al* found increased risks of health complaints and substance misuse for classes characterised by psychological abuse compared to a no-low class, and even more increased risks for classes characterised by physical abuse (Haynie et al., 2013). Goncy *et al*, also found increased risks of trauma distress for all classes vs. no-low, but patterns were less clear between classes, or for delinquency or drug use outcomes. Hébert *et al* reported that among girls there were increased risks of PTSD symptoms and of emotional distress following IPVA experiences, for the psychological & sexual victimisation class vs. no-low, and even further increased risks for the multi-victimisation class (Goncy et al., 2017). This was also the case for injuries, other psychological distress, suicidal ideation, and suicide attempts. In boys, similar patterns were observed except risks increased from no-low, to multi-victimisation, then psychological & sexual victimisation.

In summary, for younger aged samples, classes were more likely to distinguish single types of victimisation, whereas in older adolescents, they were more likely to distinguish combinations of victimisation types. There was some evidence of differential risks of negative emotional impact of victimisation experiences and health outcomes between fitted classes. There was little evidence of sex-variant class solutions, though some evidence that the relationship between classes and distal outcomes may differ between girls and boys. Few of these studies incorporated frequency of victimisation experiences.

In this study we used a bias-adjusted three-step latent class analysis to identify classes of young people according to exposure to different types and frequency of psychological, physical, and sexual victimisation by age 21, and estimate the association between these classes with negative impact and perpetration. We identified five classes: three classes that were similar for women and men were (in order of most likely classes): *No-low, Mainly psychological*, and *Psychological & physical* victimisation; two classes specific to women were: *Psychological & sexual* and *Multi-victimisation*. For both sexes, all seven types of reported negative impact were increasingly likely in this order (from *No-low* to *Multi-victimisation*). For women, all types of perpetration were most common for the *Mainly psychological, Psychological & physical* and *Multi-*victimisation classes; for men, they were most common in the *Mainly psychological* and *Psychological & physical* victimisation classes.

We can only broadly compare our findings to those from Haynie *et al*, Goncy *et al*, Sessarego *et al*, and Hébert *et al*, given the different victimisation items included, how they were captured (different questionnaire scales, and asking about different time frames of past two months to past year, compared to ‘ever’ in our study), and that perpetration was often included in models simultaneously to victimisation (Goncy et al., 2017; Haynie et al., 2013; Hebert et al., 2018; Sessarego et al., 2019). Nevertheless, we also reported a no-low and a multi-victimisation class, with similar rates of probable membership (77% and 4%, respectively) to the two studies in the older adolescents (Hebert et al., 2018; Sessarego et al., 2019), except that probabilities associated with the multi-victimisation class in Hébert *et al*’s study were noticeably higher (12% for girls, 7% for boys) (Hebert et al., 2018). We also, like previous studies, found varied risks of negative impact of victimisation experiences, including some not explored before. Unlike previous studies, we found that *psychological & sexual victimisation* and *multi-victimisation* classes were specific to females. It is possible that this is specifically the case in emerging adulthood, in a UK population, or when frequency is considered. Indeed, previous work from UK surveys found that women experience greater frequency of violence and abuse than men, and thus not taking frequency into account under-estimates the differences in experiences between the sexes (Walby & Towers, 2018). Our findings of increased negative impact for the *psychological & sexual victimisation* class compared to *no-low* in young people aged up to 21, and even further increased risk for the multi-victimisation class, correspond with what has been found previously in high-school adolescents (Goncy et al., 2017).

### Implications

There is increasing interest from researchers, service providers and policymakers, regarding IPVA among young people. IPVA prevalence is known to sharply increase during the transition from adolescence to young adulthood (Herbert et al., 2020), and young adults are a particularly understudied group in IPVA or ‘dating violence’ (Jennings et al., 2017). The reported five distinct classes of young people, who differed in their likelihood of exposure to different types of psychological, physical, and sexual IPVA victimisation, can inform the design of services dealing with young people exposed to IPVA and within future research. Psychological victimisation was a feature for all classes other than *No-low victimisation* (i.e., classes 2-5), and so identification of physical or sexual victimisation is likely to signal presence of psychological victimisation, too. This is consistent with other research on intimate partner violence (Ansara & Hindin, 2010; Bailey, Pavlou, Copas, Taylor, & Feder, 2018; Potter, Morris, Hegarty, García-Moreno, & Feder, 2020), and indicates that although young relationships takes place in a different context, for example, the individual is less likely to cohabit with their partner (Theobald, Farrington, Ttofi, & Crago, 2016), psychological victimisation can still be as pervasive.

Given the three classes that were similar between women and men, our findings support a hypothesis of gender symmetry in experiencing IPVA that involve frequent psychological and/or physical but not sexual aggression, but gender asymmetry in experiencing the most chronic IPVA victimisation or any sexual aggression (Ansara & Hindin, 2010). For young women there were two classes that were not apparent for young men: *Psychological & sexual* (class 4) and *Multi-victimisation* (class 5), both characterised by likely sexual victimisation, representing 11% of the women in the study sample.

We also provide an indication as to whether the category of IPVA types/frequency affects whether the person experiencing abuse perceives the IPVA as harmful or abusive. Each type of negative impact was least likely for the *No-*low victimisation class and the most likely for those exposed frequently to the most types of victimisation (women in the *Multi-victimisation* class [class 5] and men in the *Psychological & physical* victimisation class [class 3]) (**Figure 2**). That is, a large majority of those exposed to IPVA perceive the behaviour as harmful, and roughly in a ‘dose-response’ manner.

Our findings show a gendered relationship between victimisation and perpetration. For women, though rates were similar between the *Mainly psychological, Psychological & physical*, and *Multi-*victimisation classes, they were noticeably lower in the *Psychological & sexual* victimisation class (class 4). In contrast, in men, rates of self-reported perpetration did not vary by patterns of self-reported victimisation that were not ‘no-low’. This indicates that the mechanisms behind perpetration are more likely to vary in women. Qualitative work, that is currently in progress (http://www.bristol.ac.uk/primaryhealthcare/researchthemes/yarah-study/), will explore the possible mechanisms behind such perpetration (e.g., self-defence).

### Strengths and limitations

This study was carried out in a large contemporary cohort of young people within the UK. Though this cohort is known to over-represent white ethnic and less deprived individuals (Boyd et al., 2013), prevalence of IPVA in this cohort is similar to the wider UK general population, and other high income countries (Herbert et al., 2020; Stonard, Bowen, Lawrence, & Price, 2014; Young et al., 2018). These data have allowed us to study the relationship between victimisation patterns with negative impact and perpetration, the latter two variables rarely being available within the same dataset (Barter, 2009a; Capaldi, Knoble, Shortt, & Kim, 2012; Jennings et al., 2017; Stonard et al., 2014; Vagi et al., 2013).

We used latent class analysis to determine likely categories of victimisation types; unfortunately, indicators for goodness-of-fit were not consensual on the optimal number of victimisation classes. However, five was the largest number of classes where classes were of an acceptable size; a smaller number would increase the chance of missing important variation in victimisation responses. These five profiles were plausible given our expert knowledge of IPVA.

Previous work in this cohort indicates that the ALSPAC cohort over-represents relatively affluent, predominantly White UK populations (Boyd et al., 2013), which may limit generalisability of average probabilities of the reported five classes, though the classes properties themselves are unlikely to differ (Howe, Tilling, Galobardes, & Lawlor, 2013). At least 8% of men and 9% of women had identified as not being 100% heterosexual at age 15 (Herbert et al., 2020), but there were 25% and 30% for which this information was not available, and gender identity was not asked about by age 21. When interpreting results regarding victimisation or perpetration probabilities within each of the five classes, assumptions cannot be made about the sex or gender identity of either the person victimised or of the person who perpetrated the IPVA. In our qualitative work, we capture the experiences of socio-economically deprived individuals and ethnic, sexual orientation, and gender identity minorities, which will be used to inform as to how different experiences of different types, severity, and frequency of IPVA, and its impact might be for these groups, compared to those reported in the current study.

It must be stressed that the reported profiles use information on involvement in victimisation and perpetration, but not the relationships that this occurs in. We cannot assume that different types/severity, or total frequencies within these types, are occurring within the same intimate relationship, especially as this indicates occurrence at any time up to the age of 21. For example, it is possible that the Multi-victimisation class represents victimisation by multiple perpetrators, or that the corresponding frequent types of perpetration are being inflicted on different intimate partners. Similarly, we cannot assume temporal ordering of victimisation categories. For example, the Psychological & physical victimisation class can just as much represent psychological victimisation followed by physical victimisation, as vice versa. Our current qualitative work aims to capture such dynamics within these classes.

The negative impact questions represent participants’ *perceived* negative impact of IPVA. Perceived impact gives some indication as to whether the person being victimised perceived the behaviour as harmful or abusive, as this is not always the case (Hearn, 2013; Hester et al., 2015). Indeed, in other parts of the questionnaire over 20% of women and 10% of men reported neutral or positive impacts (e.g. ‘no effect/not bothered’, ‘thought it was funny’) (Herbert et al., 2020), and therefore though they experienced negative impacts of the abuse at the time, may not have attributed it to the events/behaviours they had been asked about. There is likely to be some error in capturing even perceived impact through these questions: if the events occurred at a young age (i.e., enough time had passed since they were asked the questions at age 21), answers to the questions may be subject to recall bias (more so than remembering whether any victimisation or perpetration ever occurred).

Perpetration is probably under-reported (Chan, 2011), which is likely to attenuate any association between IPVA profiles and perpetration type, and so the relationships described in this report (e.g., those in **Figure 3**), give a lower bound to the likely ‘true’ relationship. Future research that can combine different linked modes of reporting among young people, such as previous work that has combined survey and interview data from the same sample (Bacchus et al., 2018), can indicate to what extent this relationship can be explained by different patterns of under-reporting between victimisation classes.

## Conclusion

In this study of young people, we identified five distinct profiles of young UK people according to their IPVA victimisation patterns, including two specific to women. These profiles capture the co-occurrence of different types and frequency of IPVA and identify groups with differential risk of negative impact of victimisation and of perpetrating IPVA. Although we had limited information on sexual orientation or gender identity of those studied, the findings are consistent with emerging evidence of IPVA differentiation and its variable impact in other populations.

## Supporting information

STROBE statement

## Data Availability

ALSPAC data access is through a system of managed open access. The steps below highlight how to apply for access to ALSPAC data, including access to the Stata/R scripts used for analyses reported in this Research Article.
Please read the ALSPAC access policy, available from http://www.bristol.ac.uk/alspac/researchers/access/, which describes the process of accessing the data and samples in detail, and outlines the costs associated with doing so.

You may also find it useful to browse our fully searchable research proposals database, which lists all research projects that have been approved since April 2011.

Please submit your research proposal for consideration by the ALSPAC Executive Committee. You will receive a response within 10 working days to advise you whether your proposal has been approved.

If you have any questions about accessing data, please.

The ALSPAC data management plan describes in detail the policy regarding data sharing, which is through a system of managed open access.

## Acknowledgements

We are extremely grateful to all the families who took part in this study, the midwives for their help in recruiting them, and the whole ALSPAC team, which includes interviewers, computer and laboratory technicians, clerical workers, research scientists, volunteers, managers, receptionists and nurses.

