## Supplementary materials for "Categories of intimate partner violence and abuse (IPVA) among young women and men: Latent Class Analysis of psychological, physical, and sexual victimisation and perpetration in a UK birth cohort"

Annie Herbert^1,2^, Abigail Fraser^1,2^, Laura D. Howe^1,2^, Eszter Szilassy^1,3^, Maria Barnes^1,3^, Gene Feder^1,3^, Christine Barter^3,4^, Jon Heron^1,2^

1. Department of Population Health Sciences, University of Bristol, Bristol, UK

2. MRC Integrative Epidemiology Unit, University of Bristol, Bristol, UK

3. Centre for Academic Primary Care, University of Bristol, Bristol, UK

4. University of Central Lancashire, Preston, UK

### Box S1. Study-specific details on the three-step Latent Class Analysis

We first ran a LCA on the eight victimisation questions (six in the case of men). Variables were entered as ordinal variables (None, Once, and given numbers of responses in the more extreme categories, a combined response for A few times and Often). We ran analyses for possible solutions of two to six classes. We based our choice of optimal numbers of classes on several aspects:

- **Indicators of goodness-of-fit.** We used the Bayesian Information Criteria (BIC) and the p-values for the Bootstrap Likelihood Ratio Test (BLRT) and Vuong-Lo-Mendell Rubin (VLMR) test, to guide our decision about number of classes.(55, 56) In the case of BIC, the lowest value indicated best model fit, i.e., best fitting number of classes. A small p-value [in our case, pre-determined threshold of 0.005] for the BLRT and LMR Test, respectively, of a model for C classes indicates a better fit than a model for C-1 classes. It should be noted that in previous methodological work comparing the BIC, and BLRT and LMR Tests, the BLRT performed better than both the BIC and LMR at correctly specifying numbers of classes for LCA of categorical variables, and so when these indicators disagreed between the ‘true’ number of classes, that indicated by the BLRT would take precedent.(55) These model diagnostics are provided in **Supplementary Table S1**, which indicated very different sets of optimal classes. The BIC indicated that the best solutions would be two-class in men and five-class in women, the BLRT that this would be three-class in men and five-class in women, and the VLMR test the three-class in men and six-class in women. In this situation, we would decide to take the highest numbers of classes possible (i.e., three-class solution in men and six-class in women), as to not miss important variation in the independent variables. This is so long as it is not at the expense of face validity, a possible sex-invariant solution, or utility (each discussed below).
- **Face validity.** Co-authors on this paper (CB, ES, and GF) are experts in the field of interpersonal violence, and so given multiple indications in terms of optimal number of classes (e.g. BIC indicates one number and BLRT another), they studied the classes to determine which made more sense in terms of context and previous research findings.
- **Sex invariance.** A-priori, where possible, we attempted to establish if there were any sex-invariant classes, as this allows for comparisons of the same set of classes between females and males, particularly for purposes of studying the relationship between profiles and other variables, such as impact and perpetration. Each model diagnostic indicated different numbers of classes between sexes (**Supplementary** **Table S1**), however the classes in the three-class solution in men appear to show similar patterns to three of the five-class and six-class solutions in women (**Figure 1**).
- **Utility.** We aimed to choose a solution where classes and their probabilities would be of an adequate size size to provide stability to estimates. For example, in the six-class solution in women, several probabilities associated with this class would represent less than 5 individuals.

Entropy represents how separate or distinct classes are in terms of their features (i.e., sets of probabilities of responses), i.e., how well the model can classify individuals given these classes. The ‘best’ model can still have a low entropy given a real overlap between the ‘true’ classes. Therefore, while we didn’t base ideal number of classes on entropy, we report it for a better understanding of how well we could classify individuals based on their data. Generally, it is accepted that an entropy (which can range from 0 to 1) of at least 0.8 represents good separation. For the one-six classes solutions in men and women, entropy was at least 0.86 (ranging up to 0.91)

### Table S1: Model diagnostics between different numbers of classes

|  | **Women** | | | **Men** | | |
| --- | --- | --- | --- | --- | --- | --- |
| **Number of classes** | **BIC** | **BLRT** | **VLMR** | **BIC** | **BLRT** | **VLMR** |
| 1 | 16165.02 | N/A | N/A | 4243.19 | N/A | N/A |
| 2 | 13215.15 | < 0.0001 | <0.0001 | **3738.39** | < 0.0001 | <0.0001 |
| 3 | 12721.31 | < 0.0001 | < 0.0001 | 3769.78 | **< 0.0001** | **0.0001** |
| 4 | 12479.59 | < 0.0001 | **0.0001** | 3840.16 | 0.378 | 0.2479 |
| 5 | **12437.93** | < 0.0001 | 1.0000 | 3914.43 | 0.674 | 0.9313 |
| 6 | 12442.39 | **< 0.0001** | 0.3506 | 3989.77 | 0.386 | 0.8584 |

Values tabulated are comparing the k-class versus (k-1) class solution. Values in bold font signify that that value indicates best model fit, for that indicator.

BIC = Bayes Information Criterion; BLRT = Bootstrap Likelihood Ratio Test; VLMR= Vuong-Lo-Mendell Rubin Test; N/A = Not applicable

### Table S2. Estimated risks of negative impact types, between classes

|  | | **Women** | |  | **Men** | |
| --- | --- | --- | --- | --- | --- | --- |
| **Victimisation class** | **Impact type** | **Risk** | **Standard Error** |  | **Risk** | **Standard Error** |
| 1 (No-low) | Angry/annoyed | 1.6% | 0.004 |  | 1.3% | 0.004 |
|  | Sad | 0.2% | 0.001 |  | 0.0% | 0.000 |
|  | Upset/unhappy | 0.7% | 0.003 |  | 0.4% | 0.003 |
|  | Depressed | 0.0% | 0.000 |  | 0.0% | 0.000 |
|  | Affected work/studies | 0.0% | 0.000 |  | 0.0% | 0.000 |
|  | Anxious | 0.1% | 0.001 |  | 0.0% | 0.000 |
|  | Drank more | 0.1% | 0.002 |  | 0.1% | 0.001 |
| 2 (Psych) | Angry/annoyed | 84.5% | 0.023 |  | 83.1% | 0.036 |
|  | Sad | 86.1% | 0.059 |  | 63.7% | 0.052 |
|  | Upset/unhappy | 95.6% | 0.042 |  | 75.2% | 0.045 |
|  | Depressed | 31.4% | 0.074 |  | 13.7% | 0.038 |
|  | Affected work/studies | 27.1% | 0.064 |  | 8.2% | 0.036 |
|  | Anxious | 42.6% | 0.072 |  | 23.0% | 0.042 |
|  | Drank more | 6.8% | 0.023 |  | 9.7% | 0.025 |
| 3 (Psych & phys) | Angry/annoyed | 79.0% | 0.061 |  | 89.0% | 0.053 |
|  | Sad | 67.7% | 0.168 |  | 100.0% | 0.000 |
|  | Upset/unhappy | 81.9% | 0.127 |  | 98.1% | 0.020 |
|  | Depressed | 31.3% | 0.178 |  | 91.7% | 0.099 |
|  | Affected work/studies | 14.6% | 0.158 |  | 75.4% | 0.086 |
|  | Anxious | 40.8% | 0.172 |  | 67.2% | 0.087 |
|  | Drank more | 15.6% | 0.042 |  | 31.8% | 0.081 |
| 4 (Sex & psych) | Angry/annoyed | 94.9% | 0.023 |  |  | |
|  | Sad | 100.0% | 0.000 |  |  |  |
|  | Upset/unhappy | 98.9% | 0.011 |  |  |  |
|  | Depressed | 75.8% | 0.073 |  |  |  |
|  | Affected work/studies | 64.2% | 0.073 |  |  |  |
|  | Anxious | 83.2% | 0.066 |  |  |  |
|  | Drank more | 27.6% | 0.054 |  |  |  |
| 5 (Multi-vic) | Angry/annoyed | 76.1% | 0.051 |  |  |  |
|  | Sad | 100.0% | 0.000 |  |  |  |
|  | Upset/unhappy | 100.0% | 0.000 |  |  |  |
|  | Depressed | 90.5% | 0.050 |  |  |  |
|  | Affected work/studies | 89.5% | 0.053 |  |  |  |
|  | Anxious | 96.7% | 0.034 |  |  |  |
|  | Drank more | 51.0% | 0.074 |  |  |  |

### Table S3. Estimated risks of perpetration type*, by frequency

|  |  | **Women** | |  | **Men** | |
| --- | --- | --- | --- | --- | --- | --- |
| **Victimisation class** | **Perpetration outcome** | **Risk** | **Standard Error** |  | **Risk** | **Standard Error** |
| 1 (No-low) | Explicit psychological | 4.7% | 0.006 |  | 3.3% | 0.008 |
|  | Coercive psychological | 3.9% | 0.006 |  | 2.4% | 0.006 |
|  | Physical | 1.2% | 0.004 |  | 0.5% | 0.003 |
|  | Sexual** | 0.0% | 0.000 |  | 0.5% | 0.003 |
| 2 (Psych) | Explicit psychological | 55.8% | 0.041 |  | 68.0% | 0.048 |
|  | Coercive psychological | 39.6% | 0.038 |  | 41.4% | 0.045 |
|  | Physical | 28.7% | 0.035 |  | 11.5% | 0.027 |
|  | Sexual** | 0.4% | 0.005 |  | 9.5% | 0.025 |
| 3 (Psych & phys) | Explicit psychological | 57.1% | 0.048 |  | 50.2% | 0.095 |
|  | Coercive psychological | 43.9% | 0.047 |  | 43.6% | 0.092 |
|  | Physical | 46.8% | 0.049 |  | 11.4% | 0.059 |
|  | Sexual** | 0.8% | 0.008 |  | 5.6% | 0.042 |
| 4 (Sex & psych) | Explicit psychological | 23.7% | 0.050 |  | | |
|  | Coercive psychological | 14.2% | 0.043 |  |  |  |
|  | Physical | 11.3% | 0.040 |  |  |  |
|  | Sexual | 1.5% | 0.013 |  |  |  |
| 5 (Multi-victimisation) | Explicit psychological | 45.8% | 0.060 |  |  |  |
|  | Coercive psychological | 34.2% | 0.057 |  |  |  |
|  | Physical | 30.5% | 0.056 |  |  |  |
|  | Sexual | 3.0% | 0.022 |  |  |  |

*A response of at least ‘Once’.

**In the case of men, this represented pressured or forced into kissing, touching, something else, but not intercourse.
