## Supplementary material for "Categories of intimate partner violence and abuse (IPVA) among young women and men: Latent Class Analysis of psychological, physical, and sexual victimisation and perpetration in a UK birth cohort": STROBE statement

STROBE Statement—Checklist of items that should be included in reports of ***cross-sectional studies***

|  | Item No | Recommendation |
| --- | --- | --- |
| **Title and abstract** | 1 | *(*a) Indicate the study’s design with a commonly used term in the title or the abstract  Title: Categories of intimate partner violence and abuse (IPVA) among young women and men: Latent Class Analysis of psychological, physical, and sexual victimisation and perpetration in a UK birth cohort |
|  |  | (*b*) Provide in the abstract an informative and balanced summary of what was done and what was found  The abstract Methods and Results sections explain the data used, statistical analyses, class solution selected, probabilities of class membership, probabilities of items within classes, and distributions of impact and perpetration outcomes according to different classes. |
| Introduction | | |
| Background/rationale | 2 | Explain the scientific background and rationale for the investigation being reported  Scientific background described in Background, first and second paragraphs. Rationale described in Background, second and third paragraphs. |
| Objectives | 3 | State specific objectives, including any prespecified hypotheses  Main aim (one objective) described in Background, third paragraph (last sentence). |
| Methods | | |
| Study design | 4 | Present key elements of study design early in the paper  Methods, Participants and data collection, first paragraph: ‘We analysed data on 3,279 young people who were part of the ALSPAC (Avon Longitudinal Study of Parents and Children, formerly ‘Children of the 90s’) birth cohort study, and who had answered questions relating to IPVA (delivered in online and paper form) at 21 years old.’ |
| Setting | 5 | Describe the setting, locations, and relevant dates, including periods of recruitment, exposure, follow-up, and data collection  Setting, location, relevant dates, and data collection described in Methods, Participants and data collection, second paragraph. |
| Participants | 6 | (*a*) Give the eligibility criteria, and the sources and methods of selection of participants  Methods, Participants and data collection, first paragraph: ‘We analysed data on 3,279 young people who were part of the ALSPAC (Avon Longitudinal Study of Parents and Children, formerly ‘Children of the 90s’) birth cohort study, and who had answered questions relating to IPVA (delivered in online and paper form) at 21 years old.’ |
| Variables | 7 | Clearly define all outcomes, exposures, predictors, potential confounders, and effect modifiers. Give diagnostic criteria, if applicable.  Methods, Statistical Analyses: ‘…to determine whether impact and perpetration varied between different classes, we fitted a separate logistic regression model for different types of negative impact, and different types of perpetration as the dependent variable (outcome), respectively (all binary outcomes; in the case of perpetration outcomes, at least ‘Once’ vs. ‘Never’). Class assignment was included as an independent variable (exposure), with a correction (bias-adjustment) for potential misclassification error (Heron et al., 2015; Vermunt, 2010).  Predictors, potential confounders, and effect modifiers not relevant here. |
| Data sources/ measurement | 8* | For each variable of interest, give sources of data and details of methods of assessment (measurement). Describe comparability of assessment methods if there is more than one group  There are dedicated sections, ‘IPVA victimisation measures’, ‘Impact and IPVA victimisation’, and ‘IPVA perpetration measures’ describing how these variables were captured within the questionnaire. Table 1 provides IPVA questions (victimisation and perpetration) more explicitly). Assessment methods did not differ between participants. |
| Bias | 9 | Describe any efforts to address potential sources of bias.  Methods, Statistical Analysis: ‘Class assignment was included as an independent variable (exposure), with a correction (bias-adjustment) for potential misclassification error (Heron et al., 2015; Vermunt, 2010).’ |
| Study size | 10 | Explain how the study size was arrived at.  Methods, Participants and data collection: ‘We analysed data on the 3,279 young people who were part of the ALSPAC (Avon Longitudinal Study of Parents and Children, formerly ‘Children of the 90s’) birth cohort study, and who had answered questions relating to IPVA (delivered in online and paper form) at 21 years old.’ [Therefore, no sample size calculation was carried out.] |
| Quantitative variables | 11 | Explain how quantitative variables were handled in the analyses. If applicable, describe which groupings were chosen and why  Methods, Statistical analyses:  ‘First, we carried out an LCA where the eight victimisation questions (six for men; as ordinal variables ‘Never’, ‘Once’, ‘A few times+’) were considered.’  ‘Finally, in the third step, to determine whether impact and perpetration varied between different classes, we fitted a separate logistic regression model for different types of negative impact, and different types of perpetration as the dependent variable (outcome), respectively (all binary outcomes; in the case of perpetration outcomes, at least ‘Once’ vs. ‘Never’).’ |
| Statistical methods | 12 | (*a*) Describe all statistical methods, including those used to control for confounding  Described in a standalone section (Methods, Statistical analyses). |
|  |  | (*b*) Describe any methods used to examine subgroups and interactions  Not applicable. |
|  |  | (*c*) Explain how missing data were addressed  Not applicable. Only those who responded to the questionnaire were included, and only data from this questionnaire wave were used. |
|  |  | (*d*) If applicable, describe analytical methods taking account of sampling strategy.  Not applicable. |
|  |  | (*e*) Describe any sensitivity analyses  Not applicable. |
| Results | | |
| Participants | 13* | (a) Report numbers of individuals at each stage of study—eg numbers potentially eligible, examined for eligibility, confirmed eligible, included in the study, completing follow-up, and analysed  Methods, Participants and data collection, second and third paragraphs. |
|  |  | (b) Give reasons for non-participation at each stage  Methods, Participants and data collection: ‘Our analysis uses data from the age 21 questionnaire, which was distributed online in mid-December 2013, followed by a series of up to four reminders at three-week intervals, some of these reminders containing a paper version of the same questionnaire. 9,353 participants were sent a questionnaire (online and paper options), to which 37% (n=3,459) responded. The current study’s cohort was the 3,279 who answered questions within the IPVA section (minus one participant where sex was missing).’ |
|  |  | (c) Consider use of a flow diagram  *We do not provide one as there are only four relevant numbers to this study: mothers recruited, offspring sent questionnaire, offspring answered questionnaire, sex non-missing, and these numbers are all included in the Methods, Participants and data collection text.* |
| Descriptive data | 14* | (a) Give characteristics of study participants (eg demographic, clinical, social) and information on exposures and potential confounders  Methods, first paragraph: ‘Demographic, behavioural, and health characteristics of this sample have been described previously (Herbert et al., 2020; A. Yakubovich, Heron, Feder, Fraser, & Humphreys, 2019). Briefly, two-thirds were female, with a median age of 21 years (interquartile range: 21 to 22), the large majority were white, self-defined as heterosexual, with 92% indicating that they had been in a relationship by age 21 (Herbert et al., 2020).’ |
|  |  | (b) Indicate number of participants with missing data for each variable of interest  *Not applicable.* |
| Outcome data | 15* | Report numbers of outcome events or summary measures  Results, first paragraph: ‘Of 2,130 women and 1,149 men who responded to questionnaires at age 21, 880 (41%) and 330 (29%) reported IPVA victimisation, respectively. The most common victimisation type was psychological (35% and 26%), followed by physical (18% and 10%), then sexual (18% and 5%) (Herbert et al., 2020).’ Proportions of those with different impact and perpetration outcomes, between classes, are provided in Supplementary Tables 2 and 3. |
| Main results | 16 | (*a*) Give unadjusted estimates and, if applicable, confounder-adjusted estimates and their precision (eg, 95% confidence interval). Make clear which confounders were adjusted for and why they were included.  Unadjusted estimates of class probabilities are provided in Figure 1. Bias-adjusted risk estimates of outcomes are provided in Figures 2-3 and Supplementary Tables 2-3. |
|  |  | (*b*) Report category boundaries when continuous variables were categorized  Not applicable. |
|  |  | (*c*) If relevant, consider translating estimates of relative risk into absolute risk for a meaningful time period  Not applicable. |
| Other analyses | 17 | Report other analyses done—eg analyses of subgroups and interactions, and sensitivity analyses  Not applicable. |
| Discussion | | |
| Key results | 18 | Summarise key results with reference to study objectives  Background, third paragraph: ‘We aimed to provide a better understanding of patterns of different types, frequency, and impacts of IPVA among young men and women in the UK.’  Discussion, first paragraph: ‘We identified five classes of IPVA victimisation types and frequency, two of which only applied to women. There were differential risks of negative impact and of IPVA perpetration types, according to these classes.’ |
| Limitations | 19 | Discuss limitations of the study, taking into account sources of potential bias or imprecision. Discuss both direction and magnitude of any potential bias.  Limitations discussed under ‘Strengths and limitations’, paragraphs 2-6. Direction of bias is discussed in paragraph 6: ‘Perpetration is probably under-reported (Chan, 2011), which is likely to attenuate any association between IPVA profiles and perpetration type, and so the relationships described in this report (e.g., those in **Figure 3**), give a lower bound to the likely ‘true’ relationship.’ |
| Interpretation | 20 | Give a cautious overall interpretation of results considering objectives, limitations, multiplicity of analyses, results from similar studies, and other relevant evidence  Discussion, Conclusion: ‘In this study of young people, we identified five distinct profiles of young UK people according to their IPVA victimisation patterns, including two specific to women. These profiles capture the co-occurrence of different types and frequency of IPVA and identify groups with differential risk of negative impact of victimisation and of perpetrating IPVA. Although we had limited information on sexual orientation or gender identity of those studied, the findings are consistent with emerging evidence of IPVA differentiation and its variable impact in other populations.’ |
| Generalisability | 21 | Discuss the generalisability (external validity) of the study results  Discussion, Strengths and limitations: ‘Previous work in this cohort indicates that the ALSPAC cohort over-represents relatively affluent, predominantly White UK populations (Boyd et al., 2013), which may limit generalisability of average probabilities of the reported five classes, though the classes properties themselves are unlikely to differ (Howe, Tilling, Galobardes, & Lawlor, 2013).’ |
| Other information | | |
| Funding | 22 | Give the source of funding and the role of the funders for the present study and, if applicable, for the original study on which the present article is based  Funding statements for the current study and individual authors provided under ‘Funding Information’ section.  Conflicting interests: ‘The authors have none to declare.’ |

*Give information separately for exposed and unexposed groups.

**Note:** An Explanation and Elaboration article discusses each checklist item and gives methodological background and published examples of transparent reporting. The STROBE checklist is best used in conjunction with this article (freely available on the Web sites of PLoS Medicine at http://www.plosmedicine.org/, Annals of Internal Medicine at http://www.annals.org/, and Epidemiology at http://www.epidem.com/). Information on the STROBE Initiative is available at www.strobe-statement.org.
